# Expanded genomic surveillance and characterization of human respiratory syncytial virus infections in Minnesota, USA, 2023-2025

**DOI:** 10.1101/2025.10.10.25337732

**Authors:** Daniel Evans, Henry Kunerth, Erica Mumm, Matthew Plumb, Laura Bohnker-Voels, Sarah Bistodeau, Scott A Cunningham, Jordan Davis, Karen Martin, Kathryn Como-Sabetti, Sarah Namugenyi, Xiong Wang, Ruth Lynfield, Anna Strain, Sara Vetter

## Abstract

We report the expansion of the genomic surveillance program for human respiratory syncytial virus (HRSV) in Minnesota, USA and summarize analyses of genomes from specimens collected between July 2023 and April 2025. We continued our program into the 2024-2025 respiratory disease surveillance season, generating 2,975 high-quality HRSV genomes. Compared to those from the 2023-2024 seasons, genomes from the 2024-2025 season were collected from greater proportions of patients with HRSV-associated hospitalizations, of older age, and from a broader geographical distribution. Metrics of genetic diversity showed seasonal differences among and between HRSV-A and HRSV-B subgroups. Twenty major subclades and lineage A.D.5.4 were not observed during the 2024-2025 season. We identified 22 amino acid mutations at sites in the fusion protein associated with monoclonal antibody or receptor binding, of which 11 emerged homoplastically. We identified 39 hospitalized patients infected with HRSV strains that carried one of these mutations.

**ARTICLE SUMMARY LINE:** The Minnesota Department of Health reports key molecular and epidemiological findings after conducting genomic surveillance of human respiratory syncytial virus for two years.

## INTRODUCTION

Human respiratory syncytial virus (HRSV) continues to impose a major disease burden worldwide, causing severe pulmonary disease in populations with undeveloped or deteriorated pathogen-specific immunity (1). Methods for surveillance of HRSV have expanded in recent years, including the onboarding of prospective viral whole-genome sequencing (WGS) by national and jurisdictional health agencies and the approval of prophylactic monoclonal antibodies (MABs) and vaccines (2–4). These advances have stimulated inquiries into genetic characteristics of HRSV that enhance transmissibility and virulence, prompting the creation of WGS-based lineage classification schemes (5), studies on the prevalence of vaccine escape- and MAB resistant mutations (6), and investigations into amino acid motifs that yield competitive advantages to specific viral strains (7). These lines of effort provide insight into the molecular epidemiology of HRSV.

With the success of WGS-based surveillance of other respiratory viral pathogens (8), academic laboratories and health jurisdictions have leveraged the same technologies to gain understanding of HRSV. Recent studies on HRSV phylogenetics have advanced the field towards a standardized nomenclature that classifies viral lineages more precisely than subgrouping (5). The most recently proposed whole genome-based lineage classification scheme, which differentiates lineages based on a uniform bioinformatic protocol, has already been leveraged for recent genomic surveillance studies at the local, national, and global scales (2, 6, 7, 9, 10). The balance of legibility and precision of this classification scheme enables more efficient genomic surveillance by facilitating descriptive summaries of viral lineages over time, which can strengthen epidemiological analysis of and intervention against the circulation of HRSV (5, 11).

We previously reported on our successful initiation of genomic surveillance of HRSV infections in Minnesota, USA, for which used the novel lineage classification scheme alongside data from ongoing population-based collection of HRSV-associated outpatients, hospitalizations and deaths in the state (2). Our findings of the distributions of lineages within subgroups A and B, detection of a subclade of HRSV-B infections that demonstrated a known nirsevimab resistance-associated mutation, and cross-referencing of known infections following monoclonal antibody administration motivated us to expand our surveillance program in the subsequent respiratory viral disease surveillance season. We had three objectives: to increase the quantity and representativeness of our HRSV genomic surveillance in Minnesota, USA, to compare genetic profiles of predominating viral lineages between seasons, and to identify mutations in genomes with reported effects on resistance to monoclonal antibodies or F protein function during the HRSV infectious cycle.

## METHODS

### Collection of HRSV-positive specimens for whole-genome sequencing

The Public Health Laboratory Division of the Minnesota Department of Health recruited submission of HRSV-positive specimens from healthcare facilities and accredited clinical laboratories across the state. Institutional review board (IRB) approval was determined to be not necessary for the study of these specimens, as the surveillance of HRSV infections is a public health activity subject to Minnesota Communicable Disease Reporting Rules.

Specimens were grouped by collection date into seasons as defined by the CDC’s Respiratory Virus Hospitalization Surveillance Network (RESP-NET) (12), which reset each year on the first week that ends with a date in the month of October. This approach classified all HRSV genomes in the analysis into the 2022-2023, 2023-2024, or 2024-2025 respiratory disease surveillance seasons.

### Viral genome extraction, whole-genome sequencing, and bioinformatic analysis

HRSV genomes were extracted using the MagNA Pure 96 platform using the DNA and Viral Nucleic Acid Small Volume Kit (Roche, Germany). Extracted viral genomes were amplified by polymerase chain reaction (PCR) using 100 pairs of overlapping primers in a tiled amplicon approach, as described by Maloney et al and Dewar et al (13, 14). Amplified genomes were sequenced using either the Ligation Sequencing Kit (SQK-LSK109) with the Native Barcoding Expansion 96 (EXP-NBD196) and R9.4.1 flow cells or the Native Barcoding Kit 96 V14 (SQK-NBD114.96) and R10.4.1 flow cells on the GridION platform (Oxford Nanopore Technologies).

Genomes were assembled, assigned to viral subgroups (HRSV-A or HRSV-B), and assessed for quality using the nf-core Viralrecon pipeline (15). Genome assemblies were included in further analyses only if they achieved a sequence coverage depth of at least 40x over at least 90% of the viral genome and showed no evidence of coinfection by viral strains of both subgroups, the same thresholds employed for our previous report (2). Whole-genome lineage typing was performed using the scheme described in Goya et al, 2024 using Nextclade software (16), from multiple sequence alignments generated using Genbank reference sequences PP109421.1 (hrsv/A/England/297/2017) (HRSV-A) or OP975389.1 (hrsv/B/Australia/VIC-RCH056/2019) (HRSV-B). All HRSV genomes were typed using lineage designations refined as of April 2025.

### Phylodynamic analysis and comparisons of genetic diversity

Subgroup-specific phylogenetic trees of high-quality genome sequences were generated using the Nextstrain v9.0.0 pipeline (17). Genomes were aligned to Genbank reference sequences PP109421.1 (hrsv/A/England/297/2017, HRSV-A) or OP975389.1 (hrsv/B/Australia/VIC-RCH056/2019, HRSV-B) using Mafft v7.526 (18), and distance-scaled trees were built from those alignments using IQTree2 v2.4.0 using default settings (19).

Midpoint-rooted, time-scaled phylodynamic trees were constructed using TreeTime v0.11.4, with marginally most likely divergence dates calculated with 90% confidence intervals and polytomies resolved by stochastic subtree building (20). Pairwise single nucleotide polymorphisms (SNPs) were called from whole-genome alignments were identified using snp-dists v0.8.2 (21). Pairwise p-distances were calculated from whole-genome alignments using Mega v7.0.14 (22).

### Identification of season-specific and monoclonal antibody resistance-associated mutations

Ancestral nucleotide and amino acid sequences were inferred and translated from subgroup-specific whole-genome alignments using the ancestral sequence reconstruction and amino acid translation functions of the Augur pipeline, as integrated within Nextstrain v9.0.0 (17, 23). Genomes were searched for a list of 50 amino acid motifs potentially associated with monoclonal antibody (MAB) resistance, which was compiled from a literature review of molecular virology studies and clinical trials focused on HRSV-specific MABs and fusion (F) protein binding domains (9, 11, 24, 25, 25–30). Amino acid mutations were identified by comparing protein sequences from genomes collected in the 2023-2024 versus the 2024-2025 seasons. Mutations were inferred to have emerged during or after the summer months of 2024 if they were present only in genomes from specimens collected during the 2024-2025 season.

### Collection and analysis of epidemiological data from HRSV-associated hospitalizations

The Respiratory Syncytial Virus–Associated Hospitalization Surveillance Network (RSV-NET) conducts population-based surveillance for RSV-associated hospitalizations statewide in Minnesota (31, 32). RSV-NET cases were defined as Minnesota residents who are hospitalized with laboratory-confirmed RSV (defined as a positive laboratory test RSV during their hospitalization or during the 14 days preceding admission). Sequenced specimen data were matched with RSV-NET cases using combinations of patient first name, last name, date of birth, and specimen collection date using SAS (version 9.4; SAS Institute) and matches were manually verified. Demographic data from hospitalized patients in RSV-NET and patients with sequenced RSV specimens were then analyzed and compared.

### Statistical analyses of genomic and epidemiological data

Unpaired, two-sample Wilcoxon rank-sum tests, Fisher’s exact tests, and Kolmorogov-Smirnov tests were performed on discrete data sets of HRSV-positive patients’ age and genomes’ pairwise SNP distances using the “stats” package in R v4.5.0 software (33).

## RESULTS

### Expansion of HRSV genomic surveillance in the 2024-2025 season

Following expanded messaging from the Minnesota Department of Health aimed at clinical laboratories across the state, the number of documented unique healthcare facilities where specimens submitted for WGS were collected increased from 16 in the 2023-2024 season to 82 in the 2024-2025 season. Between October 2024 and April 2025, we generated high-quality whole-genome sequences from 2018 HRSV specimens (HRSV-A: n = 1063, 52.7%; HRSV-B: n = 955, 47.3%). Adding others from prior seasons yielded a dataset of 2975 genomes (HRSV-A: n = 1522, 51.2%; HRSV-B: n = 1453; 48.8%) [Figure 1, Supplementary Figure 1].

**Figure 1:**
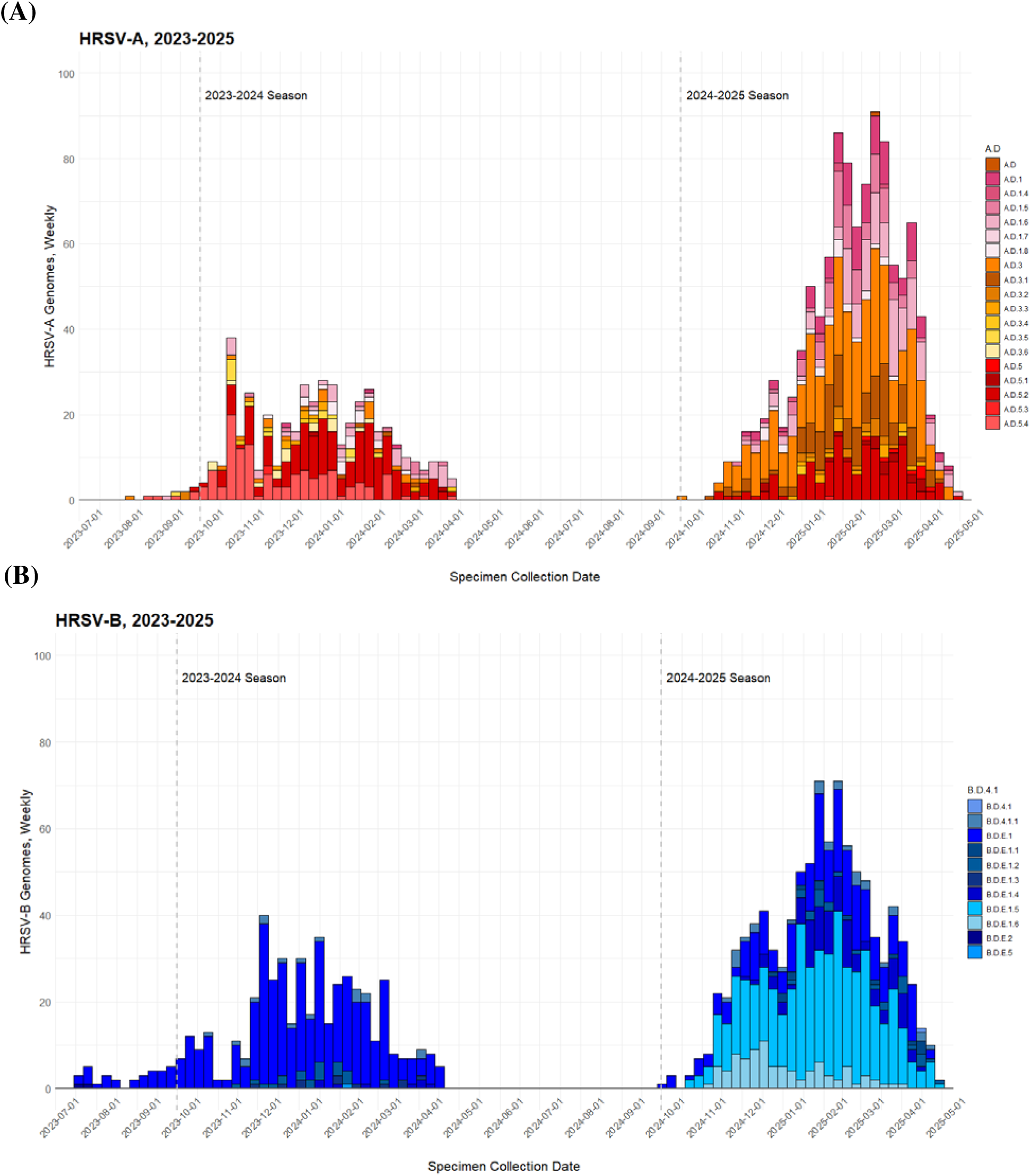
Genomic epidemiological curves of sequenced HRSV infections in Minnesota, USA, 2023-2025. The histogram shows counts of genomes by date of specimen collection, binned into 7-day periods beginning on 01 July 2023 and ending on 01 May 2023. Colors denote whole-genome lineages of sequenced genomes. Vertical dashed lines denote 01 October 2023 and 01 October 2024; Week 1 of each respiratory viral surveillance season was defined as the calendar week in which these dates fell. Panels show data for (A) HRSV-A and (B) HRSV-B genomes.

Analysis of data from submitted specimen data and the RSV-NET database of HRSV-associated hospitalizations in Minnesota showed some differences in patient characteristics between the genomic datasets of the 2023-2024 and 2024-2025 respiratory seasons. Cases from the 2024-2025 season whose specimens were successfully sequenced differed significantly in distribution of age from those from the 2023-2024 season (2023-2024: median age 1 year, interquartile range 0-3 years; 2024-2025: median age 3 years, IQR 1-29 years; Wilcoxon rank-sum test, p < 0.0001).This seasonal difference was driven by greater proportions of sequenced specimens from patients of adult age (18 years and older) in the 2024-2025 season than in the 2023-2024 season (17.9% versus 13.2% of genomes) [Table 1]. Curiously, specimens from which high-quality genomic data could not be resolved tended to be collected from patients of older age, across the entire surveillance period and in both the 2023-2024 and 2024-2025 seasons (p < 0.0001 for all time periods). Differences in proportions of subgroups A and B among genomes from the two seasons were trending but not statistically significant (Fisher’s exact test, p = 0.067).

**Table 1:** Counts and percentages of ages of patients from which HRSV specimens were collected and successfully sequenced, by 2023-2024 and 2024-2025 respiratory viral surveillance seasons.

| Age Range<br>(Years) | 2023-2024<br>Respiratory Season | 2024-2025<br>Respiratory Season |
| --- | --- | --- |
| <1 year | 285 (31.2%) | 585 (29.0%) |
| 1-4 years | 410 (44.9%) | 855 (42.4%) |
| 5-17 years | 99 (10.8%) | 216 (10.7%) |
| 18-49 years | 38 (4.2%) | 113 (5.6%) |
| 50-64 years | 19 (2.1%) | 63 (3.1%) |
| 65 years and older | 63 (6.9%) | 186 (9.2%) |
| Total | 914 | 2018 |

By reviewing demographic data from hospitalized patients with corresponding HRSV genomes – whose status was determined by their presence in the active, population-based surveillance dataset of the RSV-NET program – we observed significant differences between 2023-2024 and 2024-2025 seasons in median age, proportion of genomes from hospitalized patients, proportion of all hospitalized patients with corresponding viral genomes, and residence within the Minneapolis-St. Paul-Bloomington Metropolitan area [Table 2]. There were no statistically significant differences between these groups in proportions of subgroup A or B infections, sex of patient, or reported status of White versus other race.

**Table 2:** Genomic and epidemiological data for HRSV genomes from patients documented in the RSV-NET database for HRSV-associated inpatient hospitalizations. Statistical differences between data from the 2023-2024 and 2024-25 respiratory viral surveillance seasons were determined by Fisher’s exact tests or Wilcoxon rank-sum tests. P-values are only shown for variables with statistically significant differences by season. “N/A” = not applicable. “N.S.” = no significant difference.

|  | 2023-2024<br>and 2024-2025<br>Seasons | 2023-2024<br>Respiratory<br>Season | 2024-2025<br>Respiratory<br>Season | Statistical<br>Differences<br>By Season |
| --- | --- | --- | --- | --- |
| Sequenced<br>RSV-NET cases | 616 | 244 | 372 | N/A |
| Proportion of<br>WGS dataset (%) | 19.7% | 23.1% | 18.4% | p < 0.01 |
| Percent of<br>RSV-NET dataset | 9.1% | 7.2% | 11.1% | p < 0.001 |
| HRSV-A (%) | 314<br>(51.0%) | 118<br>(48.4%) | 196<br>(52.7%) | N.S. |
| HRSV-B (%) | 302<br>(49.0%) | 126<br>(51.6%) | 176<br>(47.3%) | N.S. |
| Female Sex (%) | 319<br>(51.8%) | 128<br>(52.5%) | 191<br>(51.3%) | N.S. |
| Median age<br>(years) | 1.9 | 1.2 | 2.5 | p < 0.0001 |
| White race (%) | 385<br>(62.5%) | 141<br>(57.8%) | 244<br>(65.6%) | N.S. |
| Minneapolis-St. Paul-Bloomington<br>Metro Area (%) | 430<br>(69.8%) | 199<br>(81.6%) | 231<br>(62.1%) | p < 0.001 |

### Changing predominance of viral lineages and genetic diversity of subgroups A and B across seasons

Comparing WGS of HRSV genomes of both subgroups A and B demonstrated that the predominance of viral lineages differed substantially between the 2023-2024 and 2024-2025 respiratory seasons. Expanding on previously reported findings (2), HRSV-A genomes from the 2023-2024 season belonged predominantly to sublineages of A.D.5 (n = 294; 65.6% of seasonal HRSV-A genomes), but sublineages of A.D.1 and A.D.3 grew to predominance among genomes from the 2024-2025 season (A.D.1: n= 364, 34.2%; A.D.3: n = 529, 49.8%). The majority of HRSV-B genomes from the 2024-2025 season belonged to sublineages that had evolved from the predominant lineage of the 2023-2024 season, B.D.E.1 (B.D.E.1.4: n = 100, 10.5%; B.D.E.1.5: n = 463, 48.5%; B.D.E.1.6: n = 90, 9.4%). Sublineage A.D.5.4, which comprised 27.2% of all HRSV-A genomes in the 2023-2024 season, was not observed in the 2024-2025 season. Phylogenetic analysis of each subgroup resolved 20 subclades of at least 10 genomes from which no descendants were observed in 2024-2025 genomic surveillance (HRSV-A: n = 8 clades, 13-127 genomes per clade; HRSV-B: n = 12 clades, 11-49 genomes per clade) and placed emerging lineages B.D.E.1.4, B.D.E.1.5, and B.D.E.1.6 in subclades separated across the HRSV-B tree [Figure 2].

**Figure 2:**
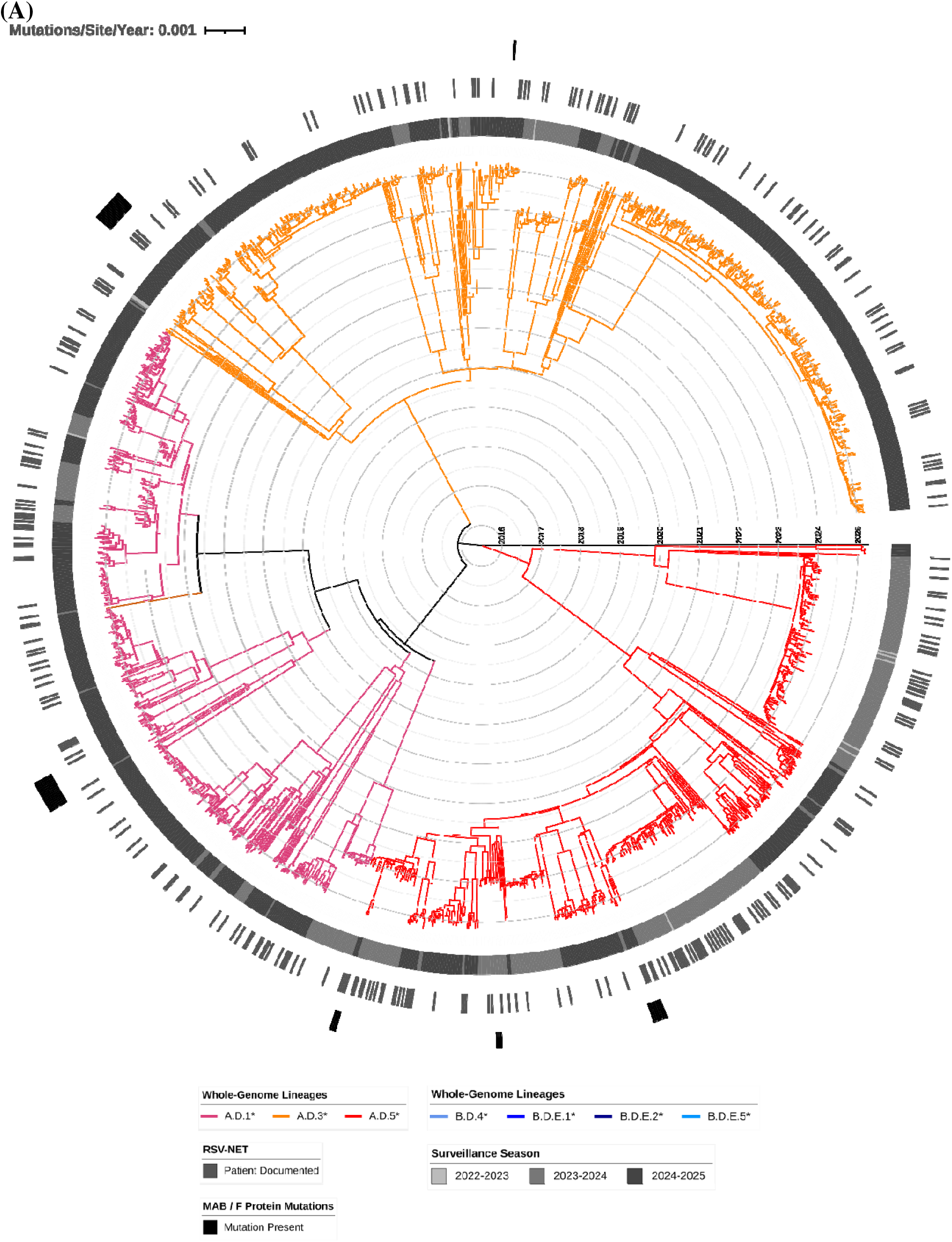
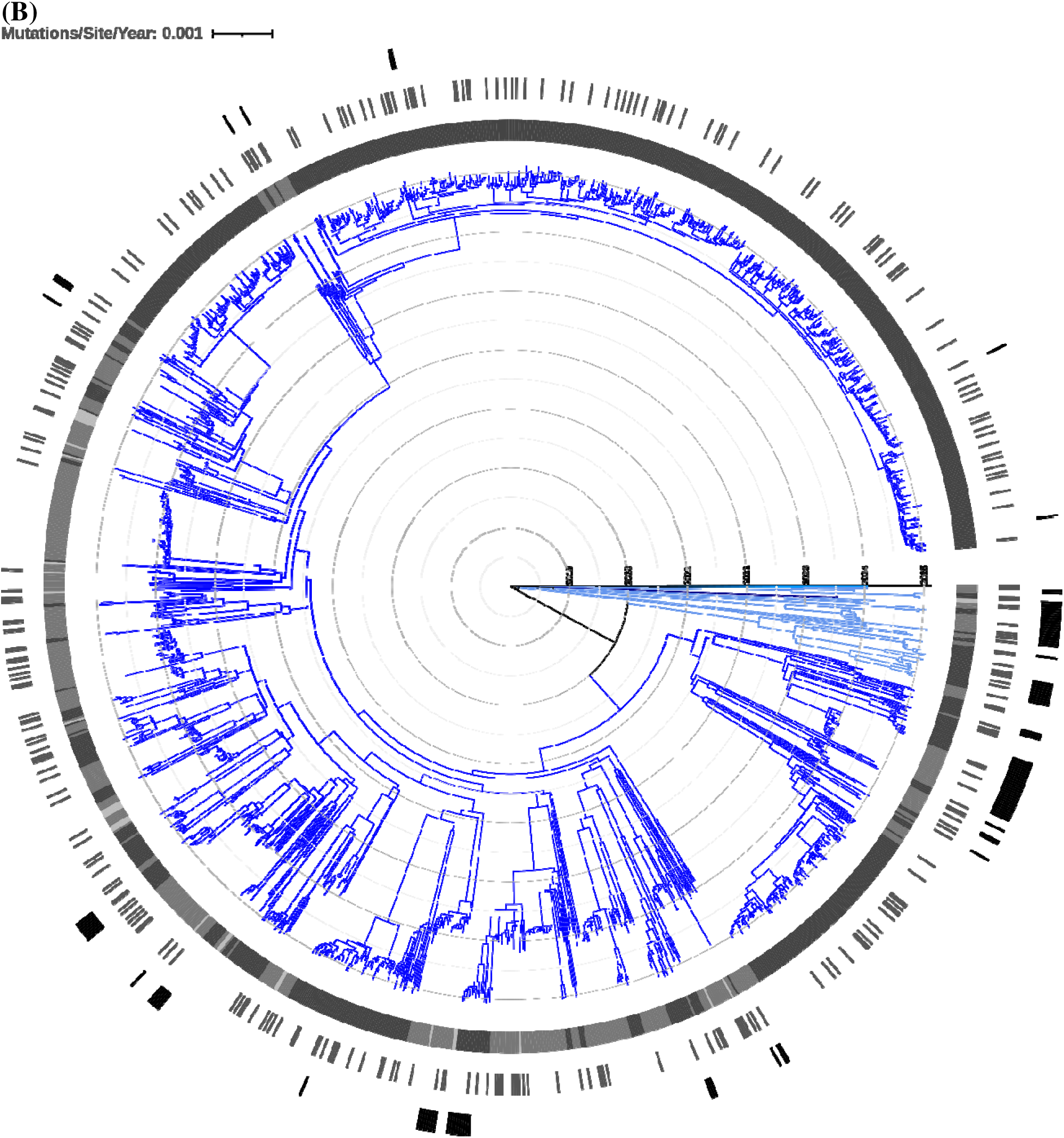
Time-scaled phylogenetic trees of (A) HRSV-A genomes and (B) HRSV-B genomes summarized in Figure 1. Major clades are colored by broad whole-genome lineage type as shown in the figure legends. Concentric circles scale each tree by time in calendar years. Annotations on each tree denote, from innermost to outermost, the respiratory viral surveillance season during which each genome was collected, the documented presence of the infected patient in the RSV-NET database of HRSV-associated inpatient hospitalizations, and the presence of at least one monoclonal antibody resistance-or F protein binding-associated amino acid mutation.

Calculations of pairwise p-distances from multiple sequence alignments of viral genomes showed that HRSV-A genomes were more genetically diverse than HRSV-B genomes across the entire genomic dataset (HRSV-A: 0.0848; HRSV-B: 0.0626), in the 2023-2024 season (HRSV-A: 0.0803; HRSV-B: 0.0536) and in the 2024-2025 season (HRSV-A: 0.0779; HRSV-B: 0.0597) seasons. Analysis of pairwise single nucleotide polymorphism (SNP) differences between genomes further showed statistically significant differences in genetic diversity between all genomes of the two subgroups (Kolmogorov-Smirnov test; D-statistic = 0.806, p < 0.001), HRSV-A genomes from 2023-2024 versus 2024-2025 (D = 0.172, p < 0.001), and HRSV-B genomes from 2023-2024 versus 2024-2025 (D = 0.432, p < 0.001) [Figure 3a]. Pairwise SNP analyses also determined that 709 (23.8%) genomes exhibited nucleotide identity to at least one other genome at 0 SNPs (HRSV-A: n = 324, 21.3%; HRSV-B: n = 385, 26.5%) and that 1227 (41.2%) genomes were separated by 1 or fewer SNPs to at least one other genome (HRSV-A: n = 596, 39.2%; HRSV-B: n = 631, 43.4% [Figure 3b]).

**Figure 3:**
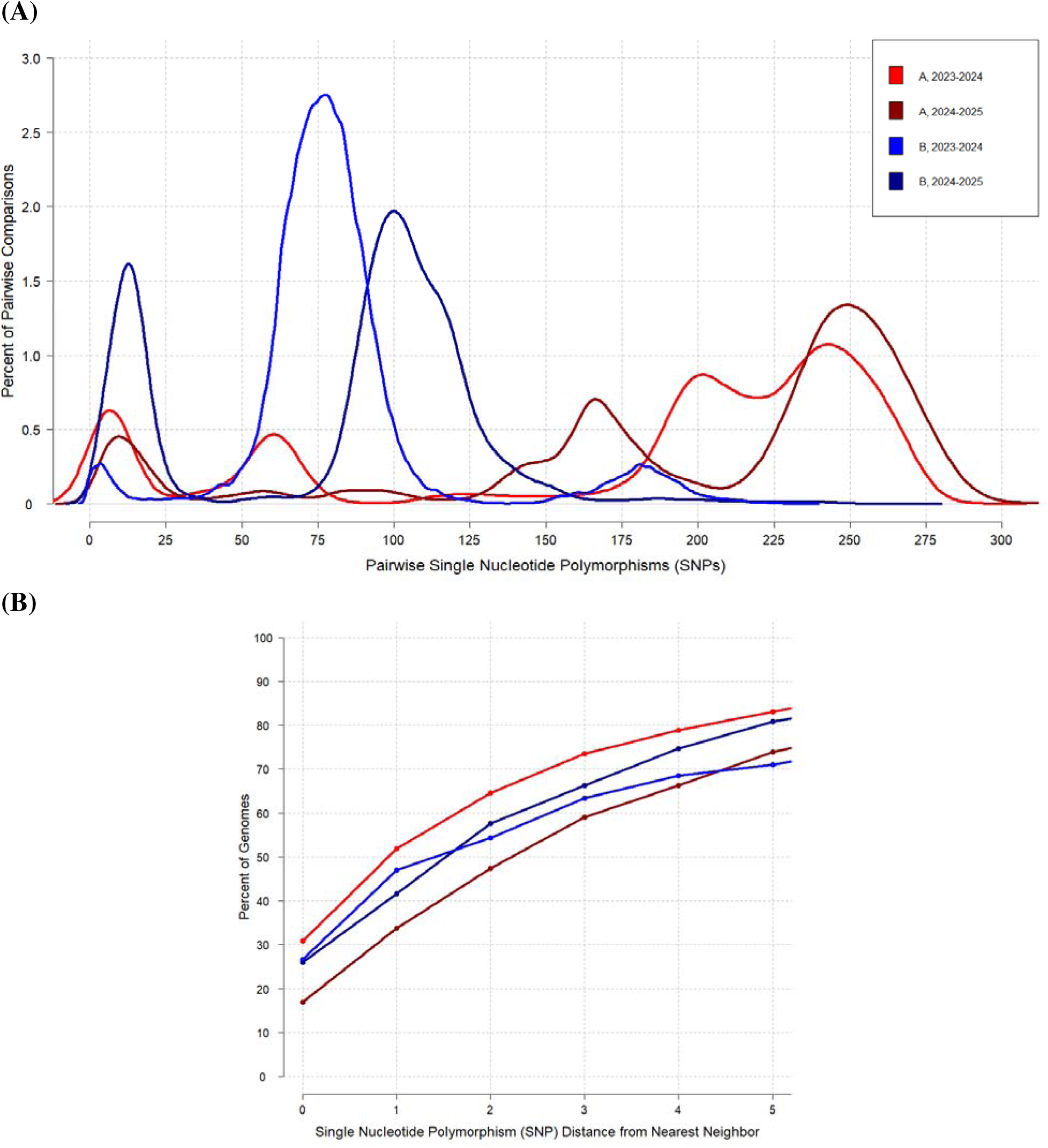
Pairwise single nucleotide polymorphism (SNP) analysis of HRSV genomes, by subgroup and respiratory season. (A) Density plots of pairwise SNP counts, scaled by percent of total comparisons between genomes. Density curves were constructed by counting SNP differences among pairs of genomes within subgroup-specific whole-genome alignments, with more closely related genomes having fewer SNPs than more distantly related genomes. (B) Cumulative distribution plots of nearest-neighbor comparisons, denoting the percent of genomes of the displayed subgroup and season that was related to at least one other genome from another patient within the displayed genetic similarity as measured by SNPs.

### Detection of emerging and homoplastic mutations linked to F protein binding and monoclonal antibody resistance

To gauge their prevalence and our surveillance program’s ability to detect MAB resistance-associated mutations, we further searched our HRSV genomes for the presence of 50 F protein substitutions – and their respective reversions – that prior literature had linked either to enhanced virus-cell fusion activity or MAB binding affinity [Table 3] (9, 11, 24–26, 28–30, 34, 35). Excluding the A103T mutation that was conserved among nearly all genomes of the A.D.5.2 lineage, 164 genomes (5.5%; RSV-A: n = 46, 3.0%; HRSV-B: n = 118, 8.1%) carried any of 22 non-synonymous mutations at 15 amino acid positions in the F protein sequence. Eleven mutations (HRSV-A: K65R, A102T, T103A, and L258S; HRSV-B: I64V, A102S, A102V, N201I, N208S, R209L, and K394R) were observed only in each subgroup’s genomes collected in the 2024-2025 season. Six mutations (K68N, K75R, A102T, K272R, T400A, and T518A) were present in single HRSV-B genomes or subclades from the 2023-2024 season but were not observed in the 2024-2025 season; these included the K68N mutation that we identified in our previous work (2). Eleven mutations emerged homoplastically in at least two distinct phylogenetic subclades of either subgroup, of which two (HRSV-A: V103A and HRSV-B: I64V) showed homoplastic emergence only in the 2024-2025 season. Within lineage A.D.5.2, the T103A reversion emerged in three of 308 (0.9%) genomes of separate subclades. The most prevalent of these homoplastic mutations was R209Q, a reversion of a known nirsevimab resistance mutation at the antibody binding site that emerged independently in 10 distinct subclades of HRSV-B (35). Nine genomes (HRSV-A: n = 1: HRSV-B: n = 8) carried two of these amino acid mutations; all six double-mutant HRSV-B genomes carried substitutions at residue 209 (R209Q: n = 5; R209L: n = 3), of which seven also carried the N211S substitution (11).

**Table 3:**
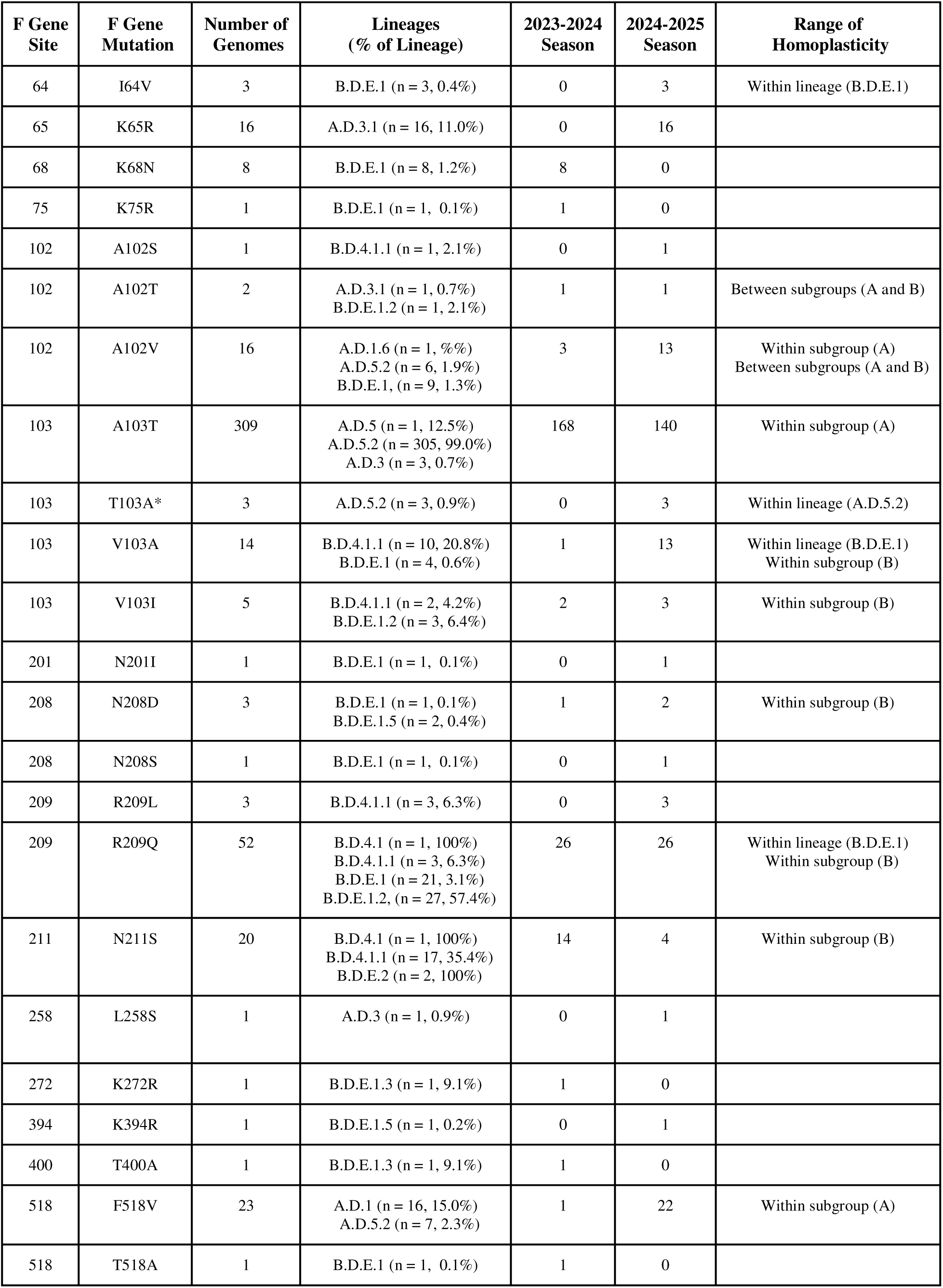
Summary of the prevalence of amino acid mutations in the HRSV F gene sequence associated with monoclonal antibody resistance or receptor binding activity. Counts and percentages of mutation presence among lineages are based on the entire genomic dataset and are not divided by respiratory viral surveillance season. Range of homoplasticity denotes whether a mutation emerged independently multiple times across the HRSV-A and HRSV-B phylogenies, and if so, the taxonomic range within which it emerged; blank cells indicate that the mutation did not emerge homoplastically.

We also reviewed epidemiological records from RSV-NET surveillance to further investigate infections by viral strains that carried any of these mutations (31, 32). Among the 616 RSV-NET patients from the 2023-2024 and 2024-2025 seasons for which we had a sequenced HRSV genome, 33 (5.4%) were immunized with nirsevimab prior to their infections. Thirty-nine (6.3%) of the 616 patients were infected with strains that carried at least one mutation reported in Table 3. One of the 33 (3.0%) hospitalized nirsevimab escape cases had viral genomes that carried at least one mutation. Four of the nine (44.4%) patients infected with double mutation-carrying viral strains were documented in RSV-NET as hospitalized.

## DISCUSSION

In this report, we summarize genomic and epidemiological results from our expanded surveillance of HRSV infections in Minnesota, USA. We also compared genetic diversity between viruses collected in the 2023-2024 versus the 2024-2025 respiratory disease surveillance seasons, highlighted seasonal differences in characteristics of HRSV-infected outpatients and inpatients, and documented mutations of potential interest that emerged in the most recent season. These findings expand upon our prior work, add context to the rapidly expanding literature on HRSV genomics, and provide insights on approaches to expand genomic surveillance of this pathogen in public health laboratories (2).

One contribution that our report provides is a comparison of genetic diversity between subgroups and seasons in a large genomic dataset from specimens collected within a constrained region. While other WGS-based studies of HRSV have compared genetic diversity between subgroups worldwide and in specific regions (3), our season-by-season comparisons of genomes from a single state provide valuable depth to the field that can contextualize findings of national-or global-level studies. It is important to note that during the genomic surveillance period, we conducted deliberate interventions to increase the quantity and representativeness of our specimen collection for the 2024-2025 season. While these efforts improved our depth of sampling across the entire state of Minnesota, they also prevented us from controlling for geographical location within the state when comparing metrics of genetic diversity. The continuous refinement of WGS-based lineage classifications for HRSV – which includes coining novel lineages as more genomes become publicly available for analysis – requires careful consideration when drawing conclusions about their emergence, predominance, and diminishment (11, 16). Furthermore, as mathematical measures of genetic diversity for subgroups A and B differed seasonally within Minnesota and cumulatively from the same metrics calculated by studies elsewhere in the world (11, 36–38), they highlight challenges of determining expected mutation rates or diversity metrics among all HRSV genomes.

Identifying and documenting MAB resistance-associated mutations has become more important in HRSV epidemiology, and WGS data from growing surveillance programs have supported these efforts (6, 39). Our detection of F protein mutations at sites known to be involved in MAB interaction or host cell fusion – including homoplastic mutations emerged independently among different subclades in the 2024-2025 season – highlights the value of more focused investigations with large quantities of sequencing data. Our detection of homoplastic mutations at amino acid site 209 in HRSV-B, a known nirsevimab binding residue, may call for further study, especially since the most prevalent mutation at this site was a reversion of a previously documented resistance mutation (35). The low prevalence of double-mutant HRSV-B genomes that carried both R209Q and mutations at the additional binding site at position 211 is consistent with findings from a newly initiated genomic surveillance program in Ireland (11).

The predominance of 209Q over 209R among our HRSV-B genomes also aligns with results from a comparison of public genomic data from 2018 versus 2024 in a recent phase I clinical trial report of another MAB candidate (40). But in contrast to that study’s additional finding of abundant mutations in position 206 that emerged between 2018 and 2024, none of the genomes in our multi-year dataset carried mutations at this position. As WGS of HRSV genomes becomes more popular worldwide (3), it is critical for these in silico observations to be evaluated through prospective clinical studies and experimental molecular virology.

An important strength of WGS-based surveillance of HRSV infections is the ability to identify numerous amino acid positions whose functions or mutations may influence the epidemiology of the virus (6). While residues in the F protein have received justifiable attention for their role in host receptor binding and MAB sensitivity, numerous laboratory and animal studies have documented other residues throughout the HRSV genome that may influence transmissibility, virulence, and host cell specificity (24). Mathematical investigations of residues under positive and negative selective pressures have also been published (3, 7), but they have not fully leveraged the rapid increase in availability of publicly available HRSV genomic data (41). The distribution of recently approved vaccines in the United States, which consist of or encode genetically modified F protein, necessitates further investigation into the emergence of mutations that reduce vaccine effectiveness (42). As additional candidate MAB immunizations are developed, tested, and licensed, the selective pressures that they impose on HRSV viral populations should also continue to be studied (4, 40). To address these important lines of inquiry, the field should continue to expand WGS-based surveillance programs for HRSV that representatively sample patients who receive MABs and vaccines and thoroughly investigate mutations that could affect the virus’s public health burden.

The successful expansion of our HRSV genomic surveillance program and subsequent discoveries were subject to several key limitations. First, although we strived to improve the representativeness of our sampling, specimen collection for WGS was voluntary for all participating laboratories and thus introduced bias into the availability of our data. Discrepancies in the distribution of our sampling between the 2023-2024 and 2024-2025 seasons also likely introduced confounding factors into our statistical comparisons of genetic and epidemiological metrics between respiratory seasons. Although we used its most recently updated version for this report, the WGS-based lineage classification schemes for HRSV-A and HRSV-B were updated several times during specimen collection and genomic analysis. Since these lineages depend on the availability of genomes and patterns of mutations within them, our summaries of lineage classifications may not perfectly reflect the true genetic diversity of HRSV worldwide. This is because HRSV genomic surveillance remains far more geographically siloed and limited in scope than similar programs for other viruses like SARS-CoV-2 and influenza (3). Our summary of MAB resistance-associated mutations was also limited by available peer-reviewed studies of HRSV, most of which were based on clinical trials of MAB candidates and molecular studies rather than from ongoing population-based surveillance initiatives (24). We also did not investigate potential vaccine escape-associated mutations, a topic of significant interest in the field given the licensure of new vaccines in the United States during our surveillance period (4, 42). Future studies should address these limitations by leveraging more representative sampling methods, identifying additional MAB resistance-associated mutations through laboratory studies of currently circulating viral lineages, mathematically calculating indicators of selective pressures on amino acid residues of potential epidemiological significance, and cross-referencing clinical records of MAB and vaccine administration prior to documented HRSV infections.

## Data Availability

All HRSV genomic data generated for this manuscript are publicly available on the National Center for Biotechnology Information website under BioProject number PRJNA1048457.

https://www.ncbi.nlm.nih.gov/bioproject/?term=PRJNA1048457

## ACKNOWLEDGEMENTS

We thank the Centers for Disease Control and Prevention’s (CDC) RSV-NET program and the Emerging Infections Program for their support of population-based surveillance of severe HRSV infections in Minnesota. We also thank D. Maloney and colleagues for their support with developing and validating laboratory methods for HRSV whole-genome sequencing. We thank all healthcare systems in Minnesota whose laboratories submitted or facilitated submission of HRSV-positive specimens for whole-genome sequencing. We also acknowledge the Minnesota Interlaboratory Microbiology Association (MIMA) for their role in encouraging HRSV-positive specimen submission.

This project was funded by the following CDC grants: Epidemiology and Laboratory Capacity (ELC) AMD Sequencing and Analytics 2 grant NU50CK000508, Minnesota Department of Health Pathogen Genomics Center of Excellence grant NU50CK000628, and Emerging Infections Program grant NU50CK000648.

---

**Supplementary Figure 1:**
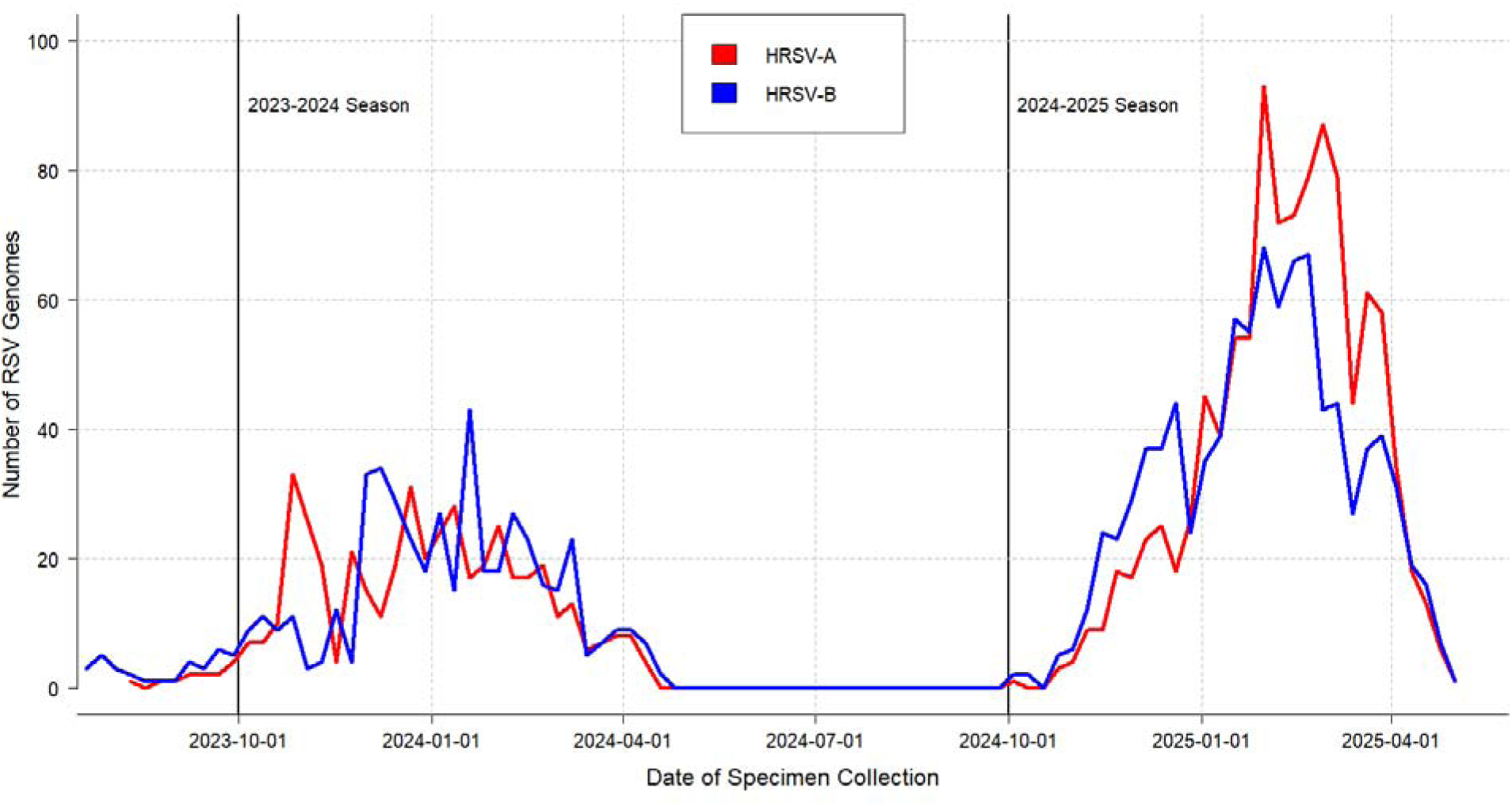
Genomic epidemiological curves of HRSV-A and HRSV-B genomes by specimen collection date across the surveillance period. Data points plotted on each curve denote counts of genomes by date of specimen collection, binned into 7-day periods beginning on 01 July 2023 and ending on 01 May 2023. Vertical solid lines denote 01 October 2023 and 01 October 2024; Week 1 of each respiratory viral surveillance season was defined as the calendar week in which these dates fell.

